# Long COVID in Cancer patients: Preponderance of Symptoms in Majority of Patients over Long Time Period

**DOI:** 10.1101/2022.07.13.22277579

**Authors:** Hiba Dagher, Anne-Marie Chaftari, Ishwaria M. Subbiah, Alexandre E. Malek, Ying Jiang, Peter Lamie, Bruno Granwehr, Teny John, Eduardo Yepez, Jovan Borjan, Cielito Reyes, Mary Flores, Fareed Khawaja, Mala Pande, Noman Ali, Raniv Rojo, Daniel D. Karp, Ray Hachem, Issam Raad

## Abstract

**Background:** An increasing number of observational studies have reported the persistence of symptoms following recovery from acute COVID-19 disease in non-cancer patients. The long-term consequences of COVID-19 are not fully understood particularly in the cancer patient population. The purpose of this study is to assess post-acute sequelae of SARS-CoV-2 infection (PASC) in cancer patients following acute COVID-19 recovery.

**Methods:** We identified cancer patients at MD Anderson Cancer Center who were diagnosed with COVID-19 disease between March 1, 2020 and Sept 1, 2020 and followed them till May 2021. To assess PASC, we collected patients reported outcomes through questionnaires that were sent to patients daily for 14 days after COVID-19 diagnosis then weekly for 3 months, and then monthly thereafter. We also reviewed patients’ electronic medical records to capture the the persistence or emergence of new COVID19-related symptoms reported during any clinic or hospital encounter beyond 30 days of the acute illness and up to 14 months.

**Results:** We included 312 cancer patients with a median age of 57 years (18-86). The majority of patients had solid tumors (75%). Of the 312 patients, 188 (60%) reported long COVID-19 symptoms with a median duration of 7 months and up to 14 months after COVID-19 diagnosis. The most common symptoms reported included fatigue (82%), sleep disturbances (78%), myalgias (67%) and gastrointestinal symptoms (61%), followed by headache, altered smell or taste, dyspnea (47%) and cough (46%). A higher number of females reported a persistence of symptoms compared to males (63% vs 37%; p=0.036). Cancer type, neutropenia, lymphocytopenia, and hospital admission during acute COVID-19 disease were comparable in both groups. Among the 188 patients with PASC, only 16 (8.5%) were readmitted for COVID-related reasons.

**Conclusions:** More than one out of two cancer patients, and more likely females, report PASC that may persist beyond 6 months and even one year. The most common symptoms are non-respiratory and consist of fatigue, sleep disturbance, myalgia and gastro-intestinal symptoms. Most of the cancer patients with PASC were managed on outpatient basis with only 8,5% requiring a COVID-19 related re-admission.

## Introduction

Clinical outcomes of COVID-19 in patients with cancer remains an area of active study with several studies showing more severe outcomes and associated risk factors such as pre-existing comorbidities, cancer stage and therapy, and immunocompromised state, compared to the non-cancer population^1-4^.

The earliest studies characterized the acute and sub-acute effects of COVID-19 on multiple organ systems^5^ while more recent work has raised concern on the chronic symptoms attributed to COVID-19 that persist beyond the expected recovery period^6,7^ with some symptoms similar to those experienced during the recovery of other viral illnesses^8-13^.

The long-term consequences of COVID-19 remain to be fully understood and there is no unequivocal consensus on the definition of post-acute sequelae of SARS-CoV-2 infection (PASC), with one adopted definition being the persistence of symptoms or new delayed complications that develop at least 4 weeks after a COVID-19 diagnosis^7,14,15^. The reported prevalence of PASC widely varies from 10% up to 87% in the general population^7-9,16-18^ and there is limited data available on PASC in cancer patients and how it affects their cancer progression, care and treatment.

Several studies suggest the persistence of symptoms past 30 days in patients with severe initial COVID-19 symptoms and those that were hospitalized due to COVID-19 ^6,17,18^, while other studies have shown a prevalence of PASC among COVID-19 outpatients as well ^8,9,19^. Since cancer patients fall in a higher COVID-19 risk group^1^ and in order to provide a better understanding of post-COVID-19 management among cancer patients, we sought to characterize the patterns of long COVID-19 in the specific cancer patient population.

## Methods

We identified patients with cancer receiving care at the University of Texas MD Anderson Cancer Center who were also diagnosed with COVID-19 disease between March 1, 2020 and Sept 1, 2020. We followed these patients as longitudinal cohort through patient-reported outcomes (PRO)-based remote symptom monitoring along with usual care with clinician visits. Patients were followed from March 2020 until May 31, 2021. Patient questionnaires were sent out remotely daily for 14 days after COVID-19 diagnosis then weekly for 3 months, and then monthly thereafter. Chart reviews were conducted for each patient’s encounter that included visits to our acute cancer care center, hospital re-admission, or clinic visit. Re-admissions were classified as either related or non-related to COVID-19 based on the reason for hospitalization as well as the reported signs and symptoms. PASC or long COVID-19 was defined as the persistence of COVID-19 related symptoms beyond 30 days of diagnosis or the emergence of new COVID19-related symptoms reported during a hospital or clinic encounter throughout the follow-up period that extended up to 14 months. COVID-19 related symptoms included among others fatigue, cough, chest tightness, dyspnea, headache, fever, altered smell or taste, myalgias, gastrointestinal symptoms (such as nausea, vomiting or diarrhea), sleep disturbance, and limitations with activities of daily living (ADLs).

### Statistical analysis

Categorical variables were compared using chi-square or Fisher’s exact test, as appropriate. Continuous variables were compared using Wilcoxon rank sum test. All the tests were 2-sided with a significance level of 0.05. The statistical analyses were performed using SAS version 9.4 (SAS Institute Inc., Cary, NC).

## Results

Among the 602 patients who were diagnosed with COVID-19 during the study period, longitudinal data was collected on 312 patients that included 188 patients who developed PASC having reported symptoms that persisted at least 30 days after COVID-19 diagnosis and 124 who did not. The remaining patients (290) could not be followed or assessed beyond 30 days. The female gender rate was significantly higher in the PASC group compared to the non-PASC group (63% vs 51%; p=0.036). The age and race were similar in both groups. The median age was 57 and 27% of both groups were above 65 years old. While the rate of hypertension was higher in the non-PASC group (56% vs 37%; p<0.001), the rate of other comorbidities such as COPD and congestive heart failures were similar in both groups. Furthermore, the type of underlying cancer was similar in both groups with the majority of patients having solid tumors. In addition, the rate of neutropenia, lymphocytopenia, lower respiratory tract infections, hypoxia, oxygen requirement, inflammatory biomarkers, COVID-19 related hospital admissions, multi-organ failure as well as medical management of COVID-19 were similar in both groups (Table 1). The 312 patients who were assessed beyond 30 days were followed for a median duration of 7 months and up to 14 months.

**Table1.** Comparing COVID-19 cancer pateints with and without long-term symptoms (at least 30 days after their COVID-19 diagnosis).

| Variables | Having PASC |  | *p*-value |
| --- | --- | --- | --- |
|  | No | Yes |  |
|  | (n=124)<br>N (%) | (n=188)<br>N (%) |  |
| Age (years), median (range) | 57 (18-85) | 57 (21-86) | 0.47 |
| Age ≥ 65 | 33 (27) | 50 (27) | > .99 |
| Gender |  |  | 0.036 |
| Male | 61 (49) | 70 (37) |  |
| Female | 63 (51) | 118 (63) |  |
| Race |  |  | 0.45 |
| Caucasian | 66 (53) | 102 (54) |  |
| Black | 24 (19) | 26 (14) |  |
| Hispanic | 28 (23) | 52 (28) |  |
| Asian | 5 (4) | 8 (4) |  |
| Other | 1 (1) | 0 (0) |  |
| Prior COPD/Bronchiolitis obliterans | 4 (3) | 7 (4) | > .99 |
| History of hypertension | 70 (56) | 69/187 (37) | < .001 |
| History of heart failure | 3 (2) | 5/186 (3) | > .99 |
| Type of cancer |  |  | 0.92 |
| Hematological malignancy | 31 (25) | 48 (26) |  |
| Solid tumor | 93 (75) | 140 (74) |  |
| Hypoxia at diagnosis | 25 (20) | 23/180 (13) | 0.08 |
| Oxygen flow at diagnosis |  |  | 0.20 |
| High flow | 6/30 (20) | 6/43 (14) |  |
| Low flow | 15/30 (50) | 15/43 (35) |  |
| None | 9/30 (30) | 22/43 (51) |  |
| Non-invasive ventilation at diagnosis | 16 (13) | 13/179 (7) | 0.10 |
| Lab values at COVID-19 diagnosis |  |  |  |
| ALC < 1 K/uL | 44/74 (59) | 57/94 (61) | 0.88 |
| ANC < 0.5 K/uL | 5/75 (7) | 5/89 (6) | > .99 |
| Hemoglobin < 10 g/dL | 23/65 (35) | 16/78 (21) | 0.06 |
| LDH ≥ 250 U/L | 29/58 (50) | 41/67 (61) | 0.21 |
| D-dimer ≥ 1 mcg/ml | 28/54 (52) | 27/61 (44) | 0.42 |
| Ferritin ≥ 500 ng/ml | 30/54 (56) | 36/59 (61) | 0.56 |
| CRP ≥ 40 mg/L | 32/56 (57) | 43/63 (68) | 0.21 |
| IL-6 ≥ 25 pg/ml | 18/41 (44) | 20/48 (42) | 0.83 |
| LRTI at diagnosis or progression to LRTI | 29 (23) | 35/182 (19) | 0.38 |
| COVID-related hospital admission | 53 (43) | 68 (36) | 0.24 |
| Remdesivir treatment | 17 (14) | 18 (10) | 0.26 |
| IL-6 pathway inhibitors - Tocilizumab | 8 (6) | 5 (3) | 0.10 |
| Convalescent plasma | 8 (6) | 15 (8) | 0.61 |
| Steroids | 18 (15) | 19 (10) | 0.24 |
| Multi-organ failure | 6/121 (5) | 8/180 (4) | 0.84 |

Note:
(Just let you know) A multivariate logistic regression analysis showed history of hypertension was the only factor that was independently associated with long-term symptoms ( $p < .001$ ), but as a protector (OR=0.43, 95% CI=0.27 to 0.69), not as a risk factor.

The most commonly reported symptoms among the 188 patients who developed PASC consisted of fatigue (82%), sleep disturbance (78%), myalgias (67%), gastro-intestinal symptoms (62%), headache (47%), altered smell and taste (47%), dyspnea (47%) and cough (46%) (Figure 1).

**Figure 1.**
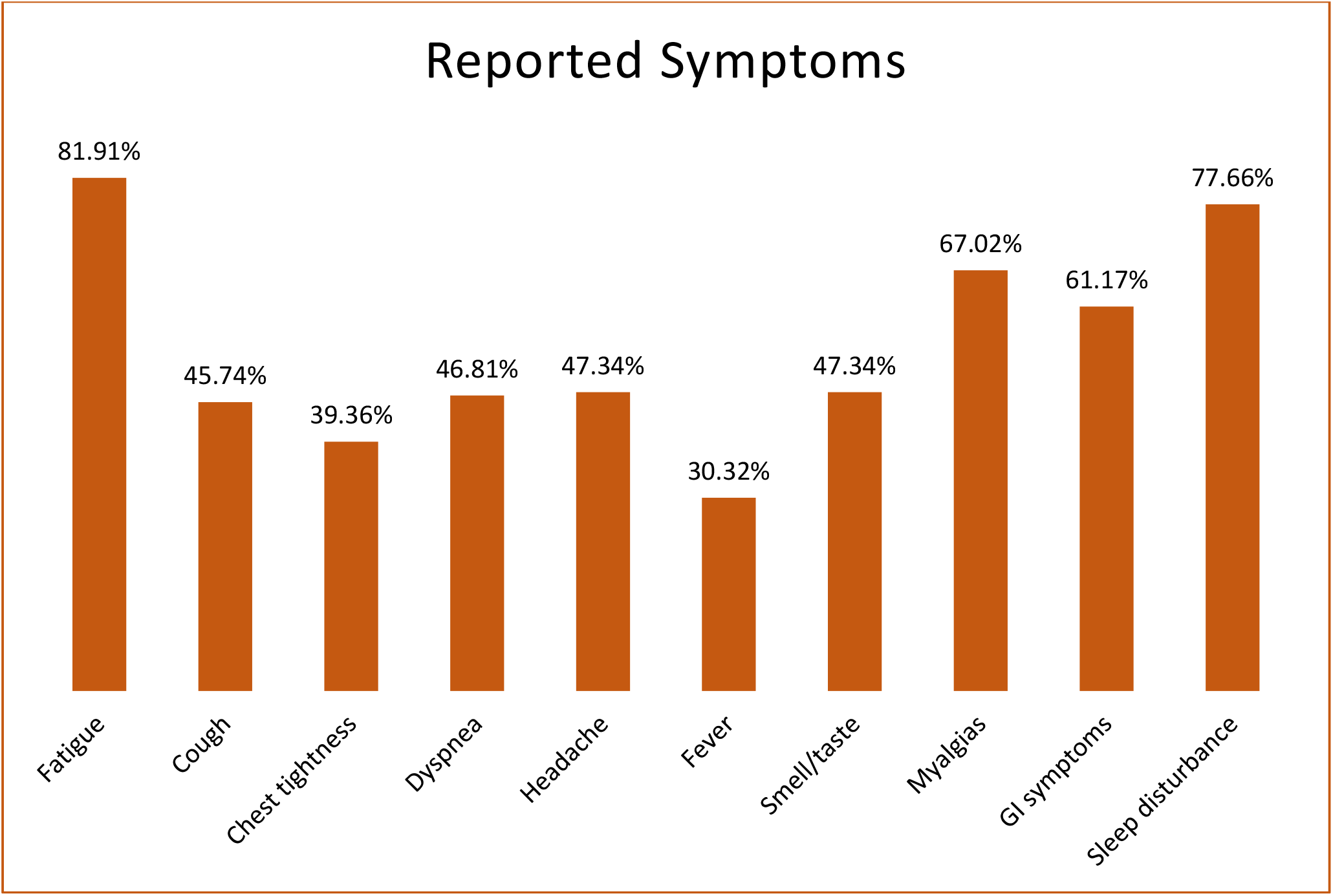
Post-Acute Sequela of SARS-CoV-2 Infection (PASC)-related symptoms (N=188).

Among the 188 cancer patients who reported PASC, 59 patients (31%) were re-admitted to the hospital during the follow-up period and beyond 30 days of acute illness, only 16 (8.5%) of the patients with PASC had a COVID-related re-admission.

## Discussion

Our data showed that 60% of 312 cancer patients diagnosed with COVID and followed up for a median duration of 7-month [up to 14-month] developed PASC. Females were more likely to report persistence of symptoms compared to males. However, cancer type, age, neutropenia, lymphocytopenia, hypoxia, severe disease, multiorgan failure, various interventions, or hospital admission during acute COVID-19 disease were not associated with higher risk of long-COVID in our study group. While hypertension was more significantly associated with no PASC during follow-up. The most common PASC symptoms were fatigue, sleep disturbances, myalgias and gastrointestinal symptoms. Furthermore, among PASC patients who were re-hospitalized for any cause during the follow-up, the admission was COVID-related in only 27%.

In a manner similar to our current study, the CDC reported that 60% of patients with immunosuppressive conditions continue to have COVID-19 symptoms beyond the acute illness^19^. In the same CDC study, hypertension was found to be significantly less often associated with PASC which is another similar finding noted in our study. It is unknown from a pathophysiologic perspective as to why hypertension, which is a risk factor for acute and severe COVID-19 should be less associated with PASC. However given the fact that the pathogenesis of acute COVID-19 is related to the interaction of the viral spike protein with the angiotensin-converting enzyme 2 (ACE 2), one could postulate that this pathway does not play an important role in long COVID compared to the residual inflammatory processes that seem to continue to occur in PASC^7,20^. Similarly, males are at greater risk of acute complications and death associated with acute COVID-19 possibly related to higher ACE 2 expression levels in the male gender^20-22^. However, in our study females were more likely to develop PASC also suggesting a different mechanism of action for PASC besides ACE 2 expression^7^.

The existing literature has also shown a higher prevalence of PASC and a longer time to recovery among those with more severe COVID-19 who required hospital admission and medical interventions^6,17,18^.However there has been several reports of PASC among patients with self-reported COVID-19 who had mild acute illness and were never hospitalized^8,9,19,23^. In our study, severe acute COVID associated with hypoxia, multiorgan failure, hospital admission or underlying risk factors for the progression of acute COVID-19 in cancer patients such as neutropenia, lymphocytopenia, hematologic malignancy or older age were not associated with increased risk of long COVID.

In the general population and following acute COVID-19, PASC symptoms occurred at a variable rate ranging from 10% to 87%^7-9,16-18^. In a manner similar to our results, fatigue was the most prevalent PASC symptom in many reports occurring in up to 87% of COVID-19 patients compared to 82% in our patient population^7,9,17,18^. Other common PASC symptoms reported in our study such as sleep disturbances, myalgias, gastrointestinal symptoms, headache, loss of smell and taste, dyspnea and cough were also widely and commonly reported in the COVID-19 literature^7-9,16-18^. However, all of these symptoms commonly occur in patients with underlying malignancy who continue to receive treatment with conventional chemotherapy, radiotherapy and immunotherapy (including checkpoint inhibitors). Hence, the rates of some of these PASC symptoms in the cancer patient population could be exaggerated and partially related to the underlying malignancy and its treatment.

In our cancer patient population, PASC was not associated with a high rate of COVID 19 related hospital admissions during the follow-up. All cause admissions occurred at a rate of 31% among our cancer patients with PASC and among those who were admitted only 27% were admitted because of COVID-19 related reasons. Hence, among all PASC patients in our study only 8.5% were admitted for COVID 19 related reasons. Furthermore, some of those few that were admitted for COVID-19 related reasons could have had reinfection with a new variant during the prolonged follow-up period or reactivation related to their immunosuppressed status. Hence, even in our high risk cancer patient population most of the symptoms associated with PASC were managed for the most part on an outpatient basis without the need for hospital admission.

Our study has its limitations. First, the retrospective nature of this study limits any firm conclusions about factors directly contributing to PASC in our COVID-19 cancer patients. Second, this is a single center study which could also limit the generalization of our results. Third, the subjective nature of surveys in the absence of quality of life assessment scales and other verified objective scales to assess reported PASC criteria could also limit our conclusions. Finally, the presence of several confounding causes of hospital admissions and overlapping symptoms in cancer patients that may be due to their underlying disease, aggressive treatments and comorbidities present their own challenge in identifying PASC among cancer patients post their COVID-19 diagnosis.

In conclusion, long COVID occurred in the majority of our cancer patients diagnosed with acute COVID-19 with a preponderance of symptoms (such as fatigue, sleep disturbances, and gastrointestinal symptoms) over a long time period. Most of the patient’s with long COVID/PASC were fairly and adequately managed on outpatient basis without the need for hospital admission. Besides the female gender, we found no other underlying condition or severity of illness during acute COVID-19 that would predict PASC.

## Data Availability

These are human subjects of multicenter trials and we are unable to share data because of IRB restrictions from different sites in different countries. The study protocol, statistical analysis plan, lists of deidentified individual data, generated tables and figures will be made available upon request by qualified scientific and medical researchers for legitimate research purposes. Requests should be sent to. Data will be available on request for 6 months from the date of publication. Investigators are invited to submit study proposal requests detailing research questions and hypotheses in order to receive access to these data. The software we used for data analysis is SAS version 9.3 (SAS Institute Inc., Cary, NC), and we have provided this information in Statistical analysis section of the manuscript.

## ACKNOWLEDGMENTS

We thank Ms. Salli Saxton, Department of Infectious Diseases, Infection Control and Employee Health, MD Anderson Cancer Center, Houston, for helping with the submission of the manuscript.

This research was supported by the National Institutes of Health/National Cancer Institute under award number P30CA016672, which supports MD Anderson Cancer Center’s Clinical Trials Office.

## References

1. Dai M, Liu D, Liu M, et al. Patients with Cancer Appear More Vulnerable to SARS-CoV-2: A Multicenter Study during the COVID-19 Outbreak. Cancer Discov. 2020;10(6):783–791.

2. Petrilli CM, Jones SA, Yang J, et al. Factors associated with hospital admission and critical illness among 5279 people with coronavirus disease 2019 in New York City: prospective cohort study. BMJ. 2020;369:m1966.

3. Wu Z, McGoogan JM. Characteristics of and Important Lessons From the Coronavirus Disease 2019 (COVID-19) Outbreak in China: Summary of a Report of 72314 Cases From the Chinese Center for Disease Control and Prevention. JAMA. 2020;323(13):1239–1242.

4. Zhou F, Yu T, Du R, et al. Clinical course and risk factors for mortality of adult inpatients with COVID-19 in Wuhan, China: a retrospective cohort study. Lancet. 2020;395(10229):1054–1062.

5. Gupta A, Madhavan MV, Sehgal K, et al. Extrapulmonary manifestations of COVID-19. Nat Med. 2020;26(7):1017–1032.

6. Barman MP, Rahman T, Bora K, Borgohain C. COVID-19 pandemic and its recovery time of patients in India: A pilot study. Diabetes Metab Syndr. 2020;14(5):1205–1211.

7. Nalbandian A, Sehgal K, Gupta A, et al. Post-acute COVID-19 syndrome. Nat Med. 2021;27(4):601–615.

8. Xiong Q, Xu M, Li J, et al. Clinical sequelae of COVID-19 survivors in Wuhan, China: a single-centre longitudinal study. Clin Microbiol Infect. 2021;27(1):89–95.

9. Goertz YMJ, Van Herck M, Delbressine JM, et al. Persistent symptoms 3 months after a SARS-CoV-2 infection: the post-COVID-19 syndrome? ERJ Open Res. 2020;6(4).

10. Ahmed H, Patel K, Greenwood DC, et al. Long-term clinical outcomes in survivors of severe acute respiratory syndrome and Middle East respiratory syndrome coronavirus outbreaks after hospitalisation or ICU admission: A systematic review and meta-analysis. J Rehabil Med. 2020;52(5):jrm00063.

11. Hui DS, Joynt GM, Wong KT, et al. Impact of severe acute respiratory syndrome (SARS) on pulmonary function, functional capacity and quality of life in a cohort of survivors. Thorax. 2005;60(5):401–409.

12. Kausler DH, Lichty W, Hakami MK, Freund JS. Activity duration and adult age differences in memory for activity performance. Psychol Aging. 1986;1(1):80–81.

13. Moldofsky H, Patcai J. Chronic widespread musculoskeletal pain, fatigue, depression and disordered sleep in chronic post-SARS syndrome; a case-controlled study. BMC Neurol. 2011;11:37.

14. Datta SD, Talwar A, Lee JT. A Proposed Framework and Timeline of the Spectrum of Disease Due to SARS-CoV-2 Infection: Illness Beyond Acute Infection and Public Health Implications. JAMA. 2020;324(22):2251–2252.

15. Greenhalgh T, Knight M, A’Court C, Buxton M, Husain L. Management of post-acute covid-19 in primary care. BMJ. 2020;370:m3026.

16. Bowles KH, McDonald M, Barron Y, Kennedy E, O’Connor M, Mikkelsen M. Surviving COVID-19 After Hospital Discharge: Symptom, Functional, and Adverse Outcomes of Home Health Recipients. Ann Intern Med. 2021;174(3):316–325.

17. Carfi A, Bernabei R, Landi F, Gemelli Against C-P-ACSG. Persistent Symptoms in Patients After Acute COVID-19. JAMA. 2020;324(6):603–605.

18. Halpin SJ, McIvor C, Whyatt G, et al. Postdischarge symptoms and rehabilitation needs in survivors of COVID-19 infection: A cross-sectional evaluation. J Med Virol. 2021;93(2):1013–1022.

19. Tenforde MW, Kim SS, Lindsell CJ, et al. Symptom Duration and Risk Factors for Delayed Return to Usual Health Among Outpatients with COVID-19 in a Multistate Health Care Systems Network - United States, March-June 2020. MMWR Morb Mortal Wkly Rep. 2020;69(30):993–998.

20. Kaseb AO, Mohamed YI, Malek AE, et al. The Impact of Angiotensin-Converting Enzyme 2 (ACE2) Expression on the Incidence and Severity of COVID-19 Infection. Pathogens. 2021;10(3).

21. Jin JM, Bai P, He W, et al. Gender Differences in Patients With COVID-19: Focus on Severity and Mortality. Front Public Health. 2020;8:152.

22. Viveiros A, Rasmuson J, Vu J, et al. Sex differences in COVID-19: candidate pathways, genetics of ACE2, and sex hormones. Am J Physiol Heart Circ Physiol. 2021;320(1):H296–H304.

23. Tenforde MW, Billig Rose E, Lindsell CJ, et al. Characteristics of Adult Outpatients and Inpatients with COVID-19 - 11 Academic Medical Centers, United States, March-May 2020. MMWR Morb Mortal Wkly Rep. 2020;69(26):841–846

